# Isoniazid preventive therapy adherence among HIV positive soldiers on antiretroviral therapy in Uganda

**DOI:** 10.1101/2023.06.05.23290987

**Authors:** Sabila Moses, Ezekiel Mupere, Joanita Nangendo, Fred Semitala, Saul Chemonges, Patience Muwanguzi, Achilles Katamba

## Abstract

**Background:** Uganda has a heavy double burden of tuberculosis (TB) and currently ranks among the seven highest TB affected countries globally. World Health Organization (WHO) recommends isoniazid preventive therapy (IPT) for the treatment of latent TB. However, the adherence to IPT in military settings has not been well documented in Uganda.

**Methods:** We conducted a cross-sectional study among 300 HIV-positive clients on antiretroviral therapy (ART) at Bombo Military Hospital in Uganda. Clients were cumulatively recruited to a threshold sample. Data were entered and analysed using Epidata client v4.6.0.6 and Stata 14.0.

**Results:** Of the 300 clients the prevalence of IPT was 94.7% CI (92.1-97.2); adherence to IPT was associated with being: aged ≥ 50 years prevalence ratio (PR) of 1.061 and a confidence interval (CI) of 95% (1.01-1.12); married [PR: 1.438, CI: 95% (1.12-1.84)]; having social support [PR: 1.498, CI: 95% (1.17-1.92)] and the role this played in IPT adherence among married participants [PR: 0.817, CI: 95% (0.72-0.93)] are factors that were found to be significant.

**Conclusion:** There is need for Bombo medical facility in Uganda to emphasis on strategies to improve access, retention and adherence to ART and IPT for young adults. Secondly, advocating for social support and behavioural interventions have been identified as requirement for improving IPT adherence among HIV positive soldiers. There is need for more research on the role that social support plays to reduce social stigma associated with HIV-positive patients. The findings for this Uganda study suggest that there is need to increase adherence to IPT for married participants living with HIV and this model could also be adopted in other resource constrained and low middle income countries.

## Background

People living with (PWH) human immunodeficiency virus infection HIV(PWH) continues to grow worldwide [1]. Sub-Saharan Africa (SSA) accounts for 71% of the global burden of HIV infections [2].

The high disease burden of HIV and tuberculosis (HIV/TB) co-infection, accounts for 95% of global TB infections [3].

Antiretroviral therapy (ART) has been documented to dramatically lower the chances of developing TB [4]. However, people with HIV still have a considerable risk of acquiring TB [5]. Combined therapies of isoniazid preventative therapy (IPT) and ART is known to reduce the burden of TB in PWH [6]. However, there is a perception that people identified in this defined cohort may be challenged with adhering to both the treatment regimens for several reasons including long duration of therapy, high pill burden and drug reaction [7].

The stipulated Ministry of Health (MOH) of Uganda guidelines on HIV management recommends IPT prophylaxis for HIV positive children and adults who do not have TB for six months [8].

A cross-sectional study in South Africa [9] found self-reported adherence to have an IPT of 72%, and randomized control trial of IPT adherence among HIV clients reported adherence of 88% in Uganda [10] Financial implications, inconveniences collecting medications, long queues, drug stock outs, pill burden, adverse side effects and lack of service integration have been shown to interrupt clients from sticking to (IPT) [7].

While extensive information is available on IPT adherence and factors associated in high burden, little is known about it in the military settings in Uganda. We aimed to conduct this study at Bombo General military hospital.

## Methods

### Study design and setting

A cross-sectional study of 300 HIV-positive clients on antiretroviral therapy (ART) at Bombo Military Hospital in Uganda was conducted.

The hospital has a capacity of 150 beds, serving over 15,000 in-patients annually and runs a full time TB/HIV clinic, which currently serves over 5082 HIV positive soldiers and over 4000 soldiers on IPT. Data on HIV and TB adherence at the facility is routinely captured in the central facility-operated data-base whenever a client visits the clinic.

Bombo Hospital has been serving as a referral Centre for various health units within the Uganda Peoples Defence Forces (UPDF) and the surrounding civilian communities since September 2012, also collaborates with the Makerere University Walter Reed Project to improve laboratory services in Uganda.

### Study population

Uganda government soldiers living with HIV/AIDS who were on ART and still on IPT treatment at BMH TB/HIV clinics between February and September 2022 were the study population. The inclusion criteria for participants in this study were clients who gave their informed consent, stayed within the military barracks were still on IPT.

### Sampling and sample size estimation

A consecutive sampling approach was used to enrol participants based on Kish Leslie formulae and assumed a z-score at 95%, CI=1.96, adherence 84% [11]. Using the margin of error of 5% and adjusted for 10% of non-response to estimate a sample of 182. For the factors associated with IPT adherence, we used the formula for two proportions to calculate the sample size, a standard normal value corresponding to 80% power of study male proportion of 50% and female proportion 50% in the study respectively was adopted. The proportion female and male clients who adhered to IPT at 88%. The calculated sample size was 472, however, only 300 clients were interviewed during the study due to transfers to various stations which reduced the number of available clients.

### Data collection/study procedures

A structured questionnaire was utilised to collect self-reported information from enrolled participants. The questionnaires were translated into Kiswahili and pretested 10 randomly selected clients. The data were collected by registered nurses trained on the study procedures and experienced in TB/HIV treatment services. The outcome variable was adherence to IPT defined as taking 95% of prescribed isoniazid (INH) doses in the last 28 days prior to the study (MOH, 2020). The independent variables included, deployments, counselling on adherence, follow up calls, fear of INH side effects, and knowledge of IPT, IPT pill burden, decision to take INH, social support, age, sex, marital status and education level. All data were checked for consistency and completeness, the necessary corrections were made, and it was double entered in Epidata v4.6.0.6

### Data analysis

All data were exported to Stata 14.0 version for analysis and summarised using descriptive statistics. The association between dependent and independent variables was evaluated using modified Poisson regression analysis model with robust standard errors. Variables with a p < 0.2 at bivariate analysis were considered for the multivariate model and variables with a p <0.05 were considered significantly associated with the outcome variable and were expressed as prevalence ratio (PR) and 95% confidence interval.

### Ethical considerations

This study was approved by the School of Medicine Research and Ethics committee (SOMREC), Number 2022-401. All participants gave informed consent to participant in the study.

## Results

### Participant demographics

The mean age of enrolled participants was 41.33(±SD 10.72) of whom 61.7% were male and 72.7% married (Table 1).

**Table 1:**
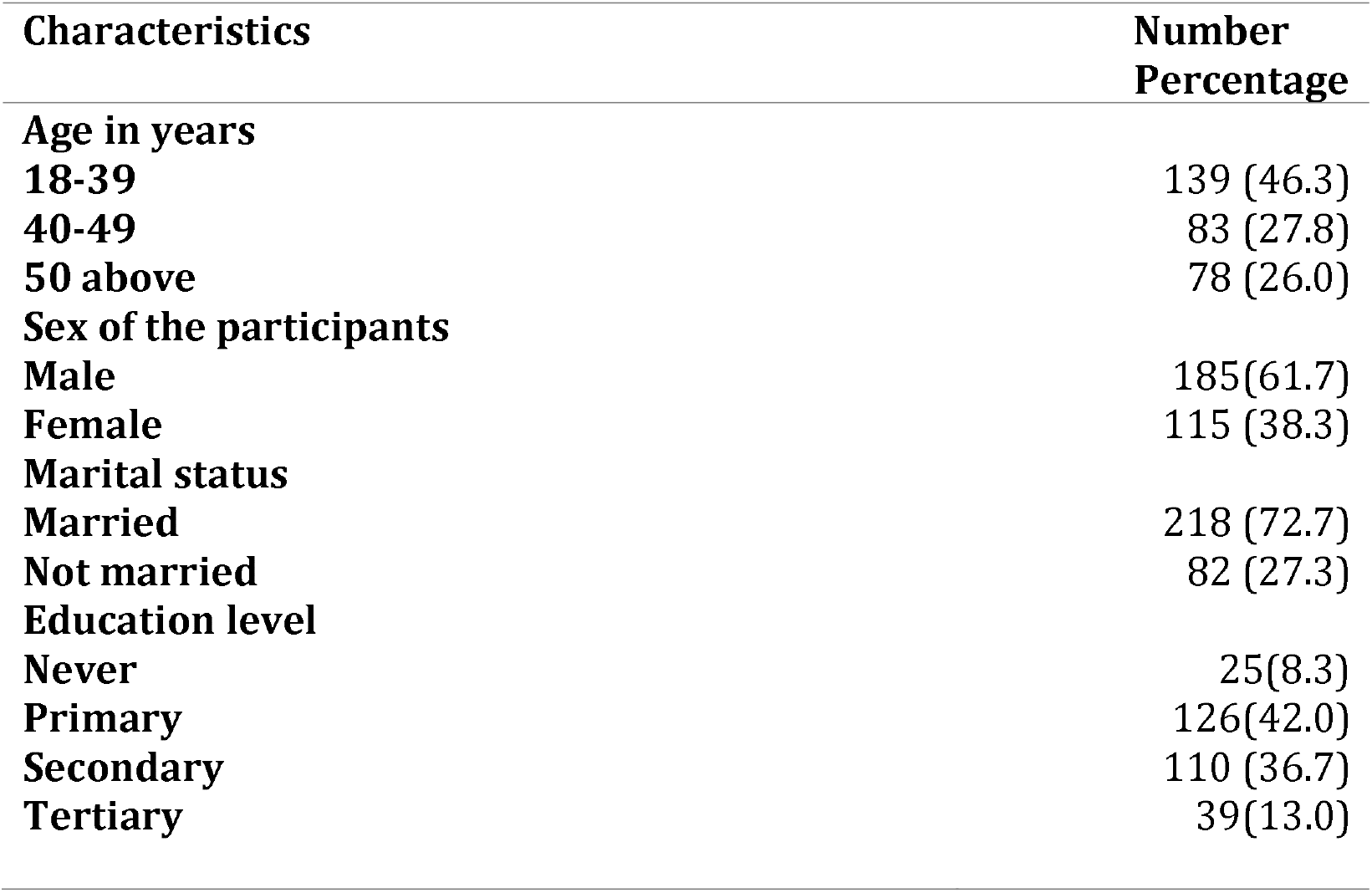
Demographic characteristics of the study of 300 soldiers on IPT at Bombo Military Hospital.

| Characteristics | Number<br>Percentage |
| --- | --- |
| **Age in years** |  |
| 18-39 | 139 (46.3) |
| 40-49 | 83 (27.8) |
| 50 above | 78 (26.0) |
| **Sex of the participants** |  |
| Male | 185 (61.7) |
| Female | 115 (38.3) |
| **Marital status** |  |
| Married | 218 (72.7) |
| Not married | 82 (27.3) |
| **Education level** |  |
| Never | 25 (8.3) |
| Primary | 126 (42.0) |
| Secondary | 110 (36.7) |
| Tertiary | 39 (13.0) |

**Table 2:**
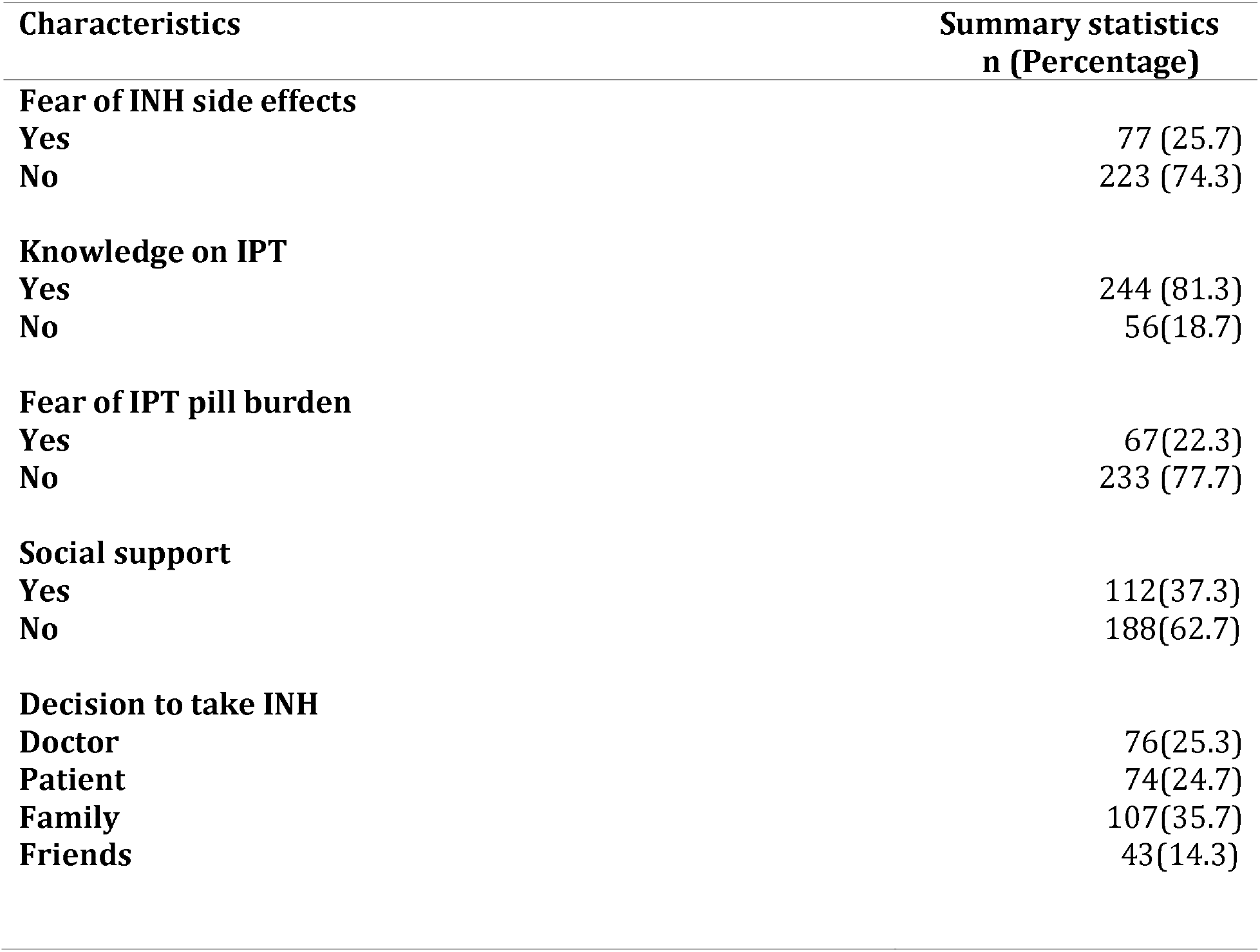
Individual personal beliefs, family and social factors of the study participants of 300 soldiers on IPT at Bombo Military Hospital.

| Characteristics | Summary statistics<br>n (Percentage) |
| --- | --- |
| **Fear of INH side effects** |  |
| Yes | 77 (25.7) |
| No | 223 (74.3) |
| **Knowledge on IPT** |  |
| Yes | 244 (81.3) |
| No | 56(18.7) |
| **Fear of IPT pill burden** |  |
| Yes | 67(22.3) |
| No | 233 (77.7) |
| **Social support** |  |
| Yes | 112(37.3) |
| No | 188(62.7) |
| **Decision to take INH** |  |
| Doctor | 76(25.3) |
| Patient | 74(24.7) |
| Family | 107(35.7) |
| Friends | 43(14.3) |

**Table 3:** Multivariate analysis of factors associated with IPT adherence among the 300 participants on ART at Bombo Military Hospital.

| Factors | PR | 95% CI | P-Value |
| --- | --- | --- | --- |
| Age in years |  |  |  |
| 18-39 | Ref | Ref |  |
| 40-49 | **0.981** | **0.91- 1.06** | 0.613 |
| 50+ | **1.061** | **1.01-1.12** | **0.030** |
| Social support |  |  |  |
| No | Ref | Ref |  |
| Yes | **1.498** | **1.17-1.92** | **0.001** |
| Marital status |  |  |  |
| No | Ref | Ref |  |
| Yes | **1.438** | **1.12-1.84** | **0.004** |
| Social support*Marital status | **0.817** | **0.72- 0.93** | **0.002** |

### Individual beliefs

Most participants (74.3%) did not fear side effects of isoniazid (INH), were knowledgeable about the IPT therapy (81.3%), and didn’t fear the pill burden (77.7%).

### Family and social factors

Most of the participants (85.3%) had not been deployed or assigned tasks in a different station during their period of treatment. A significant proportion of participants (62.7%) reported to have no social support and had the decision made by the family to starting with INH (35.7%), followed by the decision by the doctor (25.3%).

### Relationship of health workers with the clients related factors

The majority of the participating subjects reported follow up calls by the care givers (71.0%) and received counselling on adherence to IPT (86.7%).

### Prevalence of adherence and factors associated to IPT

Prevalence of self-reported adherence to IPT was 94.7%; 95% CI (92.1-97.2). Variables taken to multivariate analysis with a *p* < 0.2, included social support (*p=*0.015), dummy variable secondary education(*p*=0.186), Age 50 years and above (*p*=0.036), marital status (*p*=0.017), fear of INH side effects (*p*=0.161) and sex of the participants. The factors that were independently associated with adherence to IPT in the last 28 days, included were: 50 years and above (PR= 1.061, *P:*0.030), marital status (PR=1.438, *p*= 0.004), social support (PR=1.495, *p*=0.001) and an interaction factor between social support and marital status (PR=0.817, *p*= 0.002). Fear for INH side effects, education and sex of the participants were not statistically significant at multivariate analysis however they were significant at bivariate analysis.

## Discussion

### Prevalence of IPT adherence

In this study, it was found that 94.7% of the study participants had taken at least ≥ 95% of the prescribed isoniazid doses in the last 28 days prior to the study. The study conducted in Ethiopia, the adherence to IPT was 86.5%. This variation could be explained by the difference in the assessment of adherence. In Ethiopia, the adherence was assessed in the last 7 days, and they used random sampling opposed to consecutive sampling method which is prone to random sampling errors used in this study. This deviation could also be attributed to the way the policies of IPT uptake are enforced and implemented in the military settings compared to the civilian settings. A cross-sectional study carried out in Uganda in Butembo District on IPT found, the adherence at 84% [8]. This variation in adherence to IPT could be attributed to the difference in the cross-sectional study used in this study which is likely being affected by desirability and recall bias compared to a retrospective study. The high prevalence in this study could be explained by the following reasons: The military are good at streamlining policies and because of the discipline instilled in soldiers to obey orders, this could have facilitated the smooth implementation of IPT uptake among the soldiers. Furthermore, the use of counselling prior to medication and the information about IPT and great emphasis given to TB-HIV collaborative activity by the government, military medical leadership and implementing partners like URC. This could also be explained by enough manpower of health workers available to offer HIV services.

### Factors associated with adherence to IPT

In the multivariate analysis soldiers of age category 50+ years old were more likely to adhere to IPT. Soldiers aged 50 years and above (PR=1.061, p=0.005), were significantly associated with adherence of IPT in the last 28 days of assessment. This implied that soldiers aged 50 years and above were 1.061 times likely to adhere to IPT treatment compared to the young adults. This finding is in agreement with a study done in Tanzania which found out that soldiers aged 50 years and above were associated with IPT adherence [12], which corresponded to the current study with a tune of 98.5% (p=0.011).

It can be asserted that the elderly patients are more likely to take counselling services and advice from the health workers very serious compared to the younger adults. Additionally, in the military as a soldier approaches retirement, a soldier is assigned light duties, which gives them ample time to be with their families and adopt health-seeking behaviours. On the other hand, the poor adherence to IPT among the young adults could be due to the fact that the youthful soldiers are assigned several heavy duties both day and night in the military design. These duties are so engaging and this my hinder the soldiers from seeking medical care on time and have limited time to socialize and engage with their families. Soldiers work under orders and every activity that they undertake requires permission or a movement order which kind of policy may play a considerable big role in jeopardizing the treatment schedule of IPT. This is worse especially with the junior soldiers in the hierarchy of the army coupled with their immediate instructors who may not understand the negative impact of not taking the medicine on time and in regular basis.

The present study further revealed that having social support (PR= 1.494, *p*= 0.001) was independently associated with adherence to INH. This implies that having social support increases the likelihood adherence of IPT.

This finding of the social support is consistent with the report from a study by Munseri and colleagues [13], who reported significant influence that family and other support systems play in adherence to IPT. In their study, Munseri and colleagues further measured how likely it was for completers to have a friend or family member with TB. The finishers were also 97% more likely to have family and spouse approval for the decision to take IPT according to the study.

This finding of social support is also in agreement with reports from a study that revealed that patients rely on the support of their relatives and friends and are embarrassed to ask their jobs for permission to attend their TB treatment due to stigma [14].This is true since, in the bivariate analysis, more of the participants in this study who stuck to IPT reported receiving support from family and spouses (97.2%) than from doctors in their decision to take IPT.

The present study further revealed that being married was significantly associated with adherence (PR= 1.438, *p*= 0.004). This implies that having a partner or spouse increases the likelihood of adherence by 43.8%. This finding is consistent with a study done in South Africa in 14 public primary care clinics in Matlosana which reported 79% of the participants had a partner or a spouse as a motivator to adhere to preventive therapies compared to 21% who had no spouse [15]. This study is also in agreement with a study conducted in another part of Uganda that reported IPT uptake to be highest among married people [8], probably due to social support from the couples and the family at large to seek for IPT services at the health facility.

The study also revealed that the interaction between social support and marital status (PR= 0.817, *p*= 0.003) was associated with the adherence to IPT. This implies that additional 81.7 % of the married participants living with HIV received social support which facilitated their adherence to IPT treatment. No literature however talks about the role of social support for improved adherence to IPT among the married people living with HIV. Understanding the available social support for marrieds living with HIV reveals the important people within their social network that could be pivotal to improve their retention in care and adherence to medications and direct links between disclosure and support within families and broader community.

The role of social support among the married clients could be attributed to the fact that social support from the family takes all forms especially emotional support. Married soldiers living with HIV could have received advice and courage to adhere to their treatment through their families.

Fear for INH side effects, education and sex of the participants were not statistically significant at multivariate analysis however they were significant at bivariate analysis, which is in agreement with a study done by Munseri and colleagues which reported non-completers were more likely to cite fear of INH side effects at 14% [13].

The non-significance of the variables could be due to the small sample size, the difference in the study population, study settings, study design, sampling procedure, and difference in question comprehension.

Level of education was not significant at multivariate analysis which is not in agreement with a randomized control trial conducted in Botswana among adults which found that educated were more likely to adhere to IPT [16]. This variation could be explained by the difference in the randomized trial study design used in Botswana in contrast to cross sectional design in this study.

The gender of the participants in this study were not significant at multivariate analysis, this study differs from an Institutional-based cross sectional study conducted in 2019 in Tigari, North Ethiopia among 372 HIV positive adults, which claimed being a man was significantly associated with IPT adherence [17]. This deviation could be explained by the big sample size used in Ethiopian study as compared to the present study. The deviation could also be explained by the study population, in this study we looked at only soldiers compared to the study done in Ethiopia which looked at the general adult population.

### Limitations of the study

One limitation of this study is that adherence to IPT was measured by self-reporting. This approach is subjected to recall and social desirability biases. Secondly, a relatively small number of participants were recruited. However, the results reported herein, provide valuable insights into factors influencing adherence to IPT and highlight the value of further research in this area and HIV related factors that were not studied.

### Strength of the study

Considering that there are limited studies in this area in Uganda, the results of this study could be used as a baseline to draft program implementations to improve access, and adherence to IPT.

## Conclusion

The adherence of Isoniazid Preventive Therapy was high. The factors that were predicted to be associated with the adherence to IPT among the HIV positive soldiers on ART included, the elderly, being married, having social support and the role social support plays in the adherence of IPT among the married participants.

## Data Availability

All the data produced in this study is available upon reasonable request to the authors and contained in this manuscript

## Recommendations

The Ministry of Health could put more emphasis on strategies to improving access, retention and adherence to ART and IPT for young adult persons in Uganda. It is proposed that the army leadership can advocate for social support and behavioural interventions for improving IPT adherence among HIV positive young soldiers and the MOH should carry more research to study the role social support, plays to reduce stigma and increase adherence to IPT treatment for the married participants living with HIV in resource constrained and low middle income countries.

## Authors contribution

Sabila Moses initiated the study, performed the overall design, execution of the study, data collection and statistical analysis. Achilles Katamba and Ezekiel Mupeere supervised the study. Kalyango Joan participated in guidance throughout the project. Saul Chemonges, Patience Muwanguzi, Nangondo Joan, Fred Semitala participated in critical revision of the paper. All authors read and approved the final paper for submission.

## Acknowledgement

The authors acknowledge the data collectors, study participants, clinical epidemiology unit, Makerere university implementation science research and the administration of army leadership especially the chief of medical services-UPDF and the Director General Military Hospital, Bombo for permitting them to undertake the study. They also extend their gratitude to school of medicine research ethics committee for approving the study and Implementation science research for the financial support of the project.

